# On COVID-19-safety ranking of seats in intercontinental commercial aircrafts: *A preliminary multiphysics computational perspective*

**DOI:** 10.1101/2020.08.17.20176909

**Authors:** Prathamesh S. Desai, Nihar Sawant, Andrew Keene

## Abstract

The evolution of coronavirus disease (COVID-19) into a pandemic has severely hampered the usage of public transit systems. In a post-COVID-19 world, we may see an increased reliance on autonomous cars and personal rapid transit (PRT) systems, with inherent physical distancing, over buses, trains and aircraft for intracity, intercity, and interstate travel. However, air travel would continue to be the dominant mode of intercontinental transportation for humans. In this study, we perform a comprehensive computational analysis of typical intercontinental aircraft ventilation systems to determine the seat where environmental factors are most conducive to human comfort with regards to air quality, protection from orally or nasally released pollutants such as CO_2_ and coronavirus, and thermal comfort levels. Air velocity, temperature, and air pollutant concentration emitted from the nose/mouth of fellow travelers are considered for both Boeing and Airbus planes. In each plane, first class, business class, and economy class sections were analyzed. We present conclusions as to which is the optimum seat in each section of each plane and provide the data of the environmental conditions to support our inferences. The findings may be used by the general public to decide which seat to occupy for their next intercontinental flight. Alternatively, the commercial airliners can use such a model to plan the occupancy of the aircraft on long-duration intercontinental flights (viz., Airbus A380 and Boeing B747).

## INTRODUCTION

In the present-day world, where on one hand public policy makers are grappling to contain the spread of COVID-19 [1], on the other hand certain transit systems are facing existential threats [2]. COVID-19 has crippled public transit systems such as rails, buses, ride-shares, car rentals, and domestic flights. Lowered ridership, increased sanitation requirements in between rides, and the associated expenses can force these systems into liquidation (see this report on Hertz [3]). There is a high possibility of a leapfrog towards autonomous cars and personal rapid transit (PRT) systems for intercity, intrastate, and interstate travel. However, air travel would continue to be the dominant mode of intercontinental transportation of humans. The airliners would have to pay special attention towards indoor air quality and thermal comfort.

Numerous requirements and standards exist that dictate acceptable indoor air quality. When designing a ventilation system for an airplane fuselage (or cabin) the challenge is to balance these considerations with fuel efficiency since all air entering the cabin is being taken from the compression stage of the engine, reducing thrust [4]. The air is first conditioned to the desired temperature and humidity then injected into the cabin through vents on the ceiling at a rate which maximizes efficiency. The air circulates and then exits through an exhaust along the outside of the floor. The air is recirculated and mixed with fresh air before being injected again. Common problems with this system occur when too much recirculated air is introduced back into the cabin with too little fresh air, resulting in headaches and tiredness for passengers [5]. Another concern is the dispersion of contaminants introduced into the cabin through a couch or sneeze. Although a multitude of research has been performed to optimize the ventilation system to most effectively control these factors, implementation of new technology or equipment in the aircraft industry is notoriously slow due to stringent safety guidelines. For this reason, the authors have chosen to examine the existing system used in commercial aircrafts and perform an analysis that will allow a passenger to make an educated decision about which seat will provide the most preferable conditions.

As previously mentioned, extensive research has been performed in this area simulating the airflow through an aircraft cabin often with a focus on optimizing the system, or tracking contaminant dispersion. Additionally, large scale experiments have been performed to validate the results obtained through computational fluid dynamics (CFD) simulations. Although these works are not directly pertinent, especially when considering different ventilation simulations, a selection will be briefly discussed because many parameters of the flow and cabin and techniques for numerical modeling guided the work done in this paper. One example, work done by Zhang et al. criticizes current systems for producing a dry environment with excessive air mixing [5]. As a solution they developed an under-aisle distribution system which supplies dry air and humidified air through perforated aisles and increases humidity levels and lowers CO2 concentrations throughout the cabin. Subsequent work, also by Zhang et al. [6], investigates the viability of a personal chair armrest air delivery system that delivers fresh outside air directly to each passenger’s breathe zone. The exhausts are moved to be overhead instead of on the floor as is typical. They find that this system results in undesirable vertical temperature gradients, but is able to prevent contaminant release at any level [6].

Numerous studies have also been performed using an unsteady inlet condition. Although we chose a steady inlet condition, which is consistent with most simulations, the results from unsteady flow provide an interesting comparison. Yan et al. focused on the spreading of disease from a cough or sneeze in an aircraft cabin. Experiments were performed in full scale cabin mockup with an unsteady inlet velocity and tracer gas used to visualize the flow field. Results showed a more complex, but narrower spread than with unsteady flow. Results were compared to CFD results which showed good correlation [7]. Similarly, Wu et al. performed CFD analysis on a B767 cabin section and showed that an unsteady air supply had more desirable temperature and CO2 distribution than the equivalent steady flow situation [8].

Experiments performed using full scale models of airplane cabins are especially useful because they provide validation for our results. Liu et al. [9] provides a review of many experimental measurements and numerical simulations done to predict flow in aircraft cabin. Full scale experiments are shown to be the most reliable and accurate, however they are the most expensive and time consuming. He concludes simulations are promising as an alternative [9]. A subsequent study [10] by the same group used laser tracking and reverse engineering to generate an accurate model of cabin geometry and measurements of boundary conditions (at diffusers) and flow fields. They concluded that flow and boundary conditions in real cabins were complex and velocity and turbulence of inlet flow varied significantly from one opening to the next. Hence, an averaged value of this data serves as a valid approximation for simulations [10].

Wang et al. [11] acquired experimental data from a full scale mock-up of a Boeing cabin with 35 mannequins including heaters in their body sections. The goal was to evaluate ventilation effectiveness and characterizing air distribution. This study produced results similar to previously published accepted data and proved that modeling passenger heat emittance from just the torso section is a reasonable simplification [11]. A research group lead by Garner [12] performed a study concerned with an injection of particles to the ventilation system mid-flight in relation to a terrorist threat. They measured the velocity field at various points in the actual plane at operating conditions and perform 2D CFD analysis to compare. CFD results match nicely with experimental data suggesting that a 2D computational model is a simplification that does not jeopardize the accuracy of the solution [12].

Bosbach et al. [13] used particle image velocimetry in a full scale mock-up of an aircraft cabin to get velocity field data throughout the domain in order to examine relative effects of forced and natural convection in the temperature distribution in the cabin. This was done in order to validate a CFD analysis showing these effects that was created to minimize computational time. These results gave us confidence that the accuracy of our results would be maintained if natural convection was ignored [13].

Airliners offering intercontinental flights will have to rethink seat placements to ensure safe and uniform environment for each traveler. One option is to permanently remove less suited seats, while the other is to seat passengers only on the better positioned seats. A few recent studies [14–18] have tried to numerically model the indoor spread of COVID-19 viral particles using CFD. A couple of studies [19,20] have explicitly studied the transport of pollutants inside aircraft cabins. However, the flow physics of virus laded fluid is not trivial [21]. Additionally, no study has yet modeled the airflow, transport of nasally and orally released pollutants like CO2 and coronavirus, and thermal efficiency inside two of the most common long-duration intercontinental aircrafts (viz., Airbus A380 and Boeing B747). The authors of the current study present a preliminary computational model to do the aforementioned and rank the seats of the first class, business class, and economy class cabins inside A380 and B747.

## PROBLEM SETUP

Problem physics included solving for the air flow, temperature distribution, and CO2 transport inside the airplane cabin. CO2 was chosen as a representative pollutant that is released orally or nasally.

As the geometry dimensions were very large and the inlet velocity of air was very low, the flow was expected to be nearly incompressible. Therefore, it was modeled using incompressible Navier-Stokes equations.

Continuity equation: *∇.**V*** = 0

Momentum conservation: 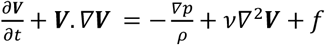

Temperature distribution was modeled using the energy conservation equation:

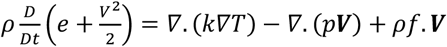

CO_2_ transport was modeled using the mass transport equation:

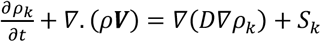

Where,

***V*** – Fluid flow field velocity, in m/s
*p* – Fluid flow field pressure, in N/m^2^
*v* – Fluid kinematic viscosity, in m^2^/s
*f* – Body force acceleration, in m/s^2^
*_ρk_* Mass concentration of species k, in kg/m^3^
*D* – Molecular diffusion coefficient
*S_k_* – Source term for species k, e.g. due to chemical reactions

This paper modeled 2D sections for the airplane cabins, so it was important to select the right section to correctly represent the 3D problem. The initial section was across the head and torso of a human, as shown in Fig. 1. But human bodies occupied minimum space of the cabin volume hence it was not the correct representation the 3D space. Additionally, the human body model interfered with important flow regions, thus stopping the crucial flow patterns from developing. As a result of this, a new section, passing through the laps of human and airplane seat was selected as seen in Fig. 2.

**Figure 1.**
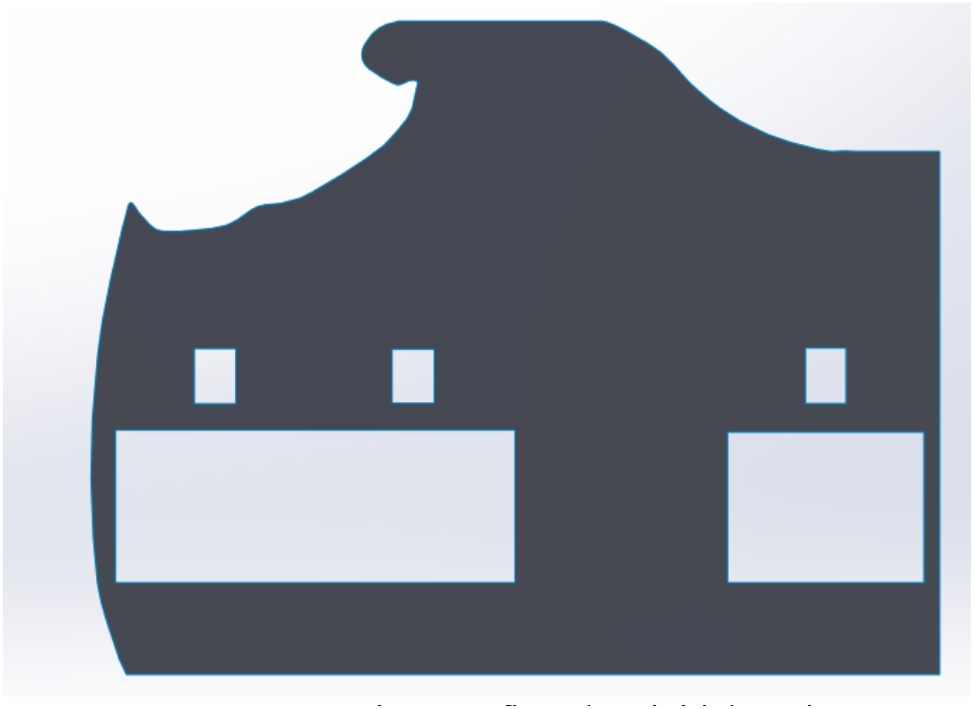
Boeing 747 first class initial section

**Figure 2.**
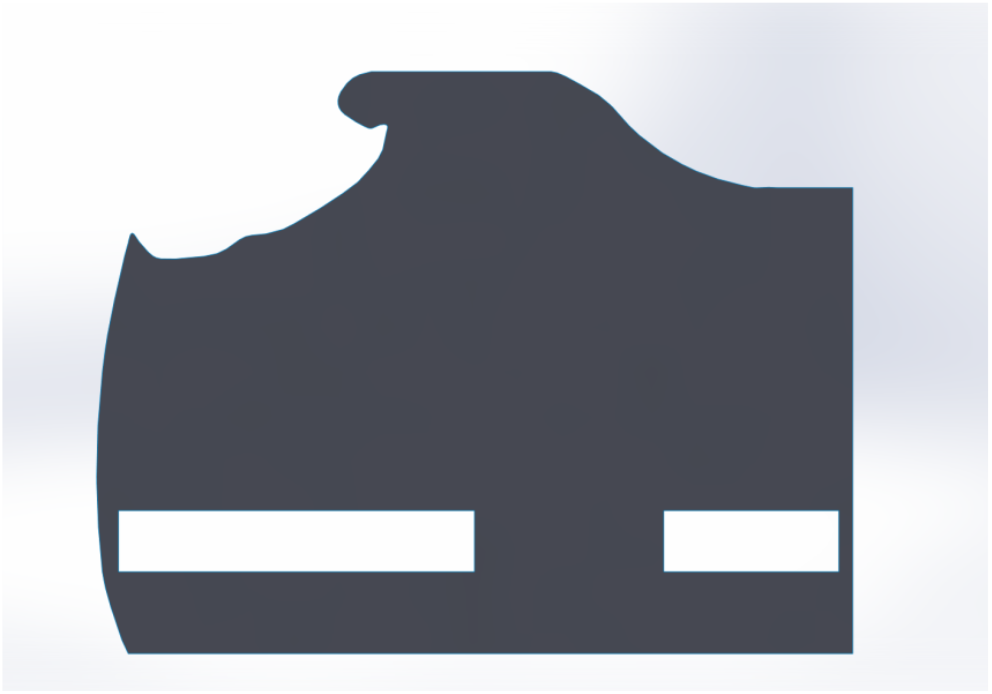
Boeing 747 first class final section

Wang et al [11] mentioned that most of the air supplied from one passenger row is circulated and finally exhausted in the same row. Also, Yan et al [7] mentioned in their paper, that most pollutants are transported only within the releasing half and seldom cross the middle line. Therefore, the authors have assumed symmetry in computational domain and hence have considered only half section of the plane.

Boundary conditions for the problem are represented in Fig. 3 and Table 1 as shown below.

**Figure 3-a.**
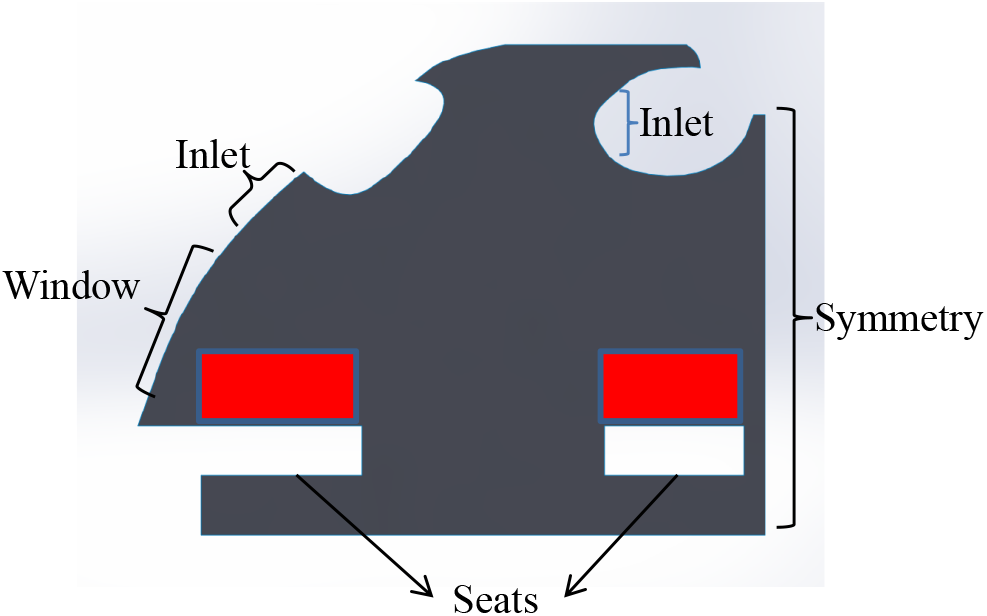
Airbus A380 first class boundary conditions. The red blocks represent the human bodies and the associated heat sources.

**Figure 3-b.**
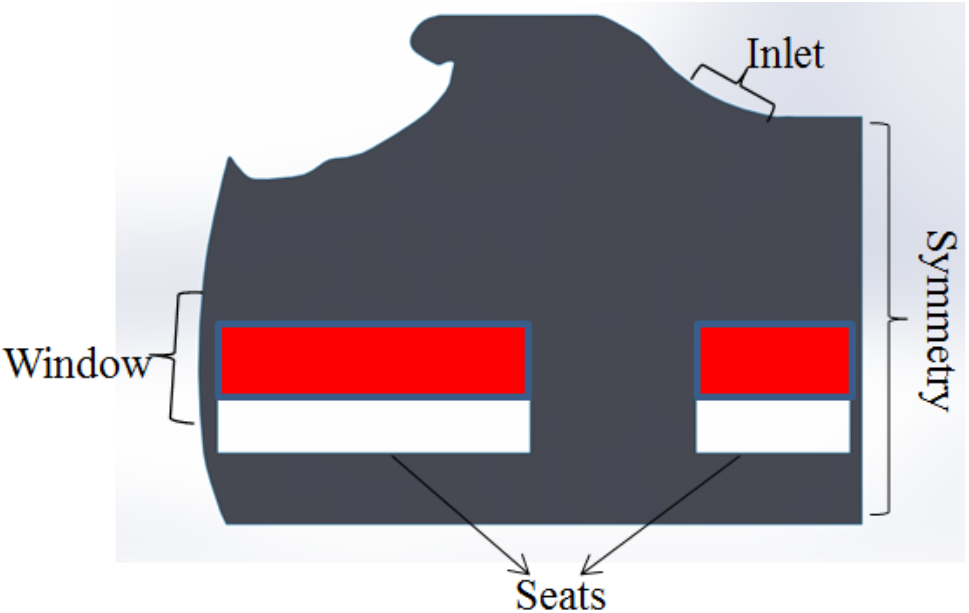
Boeing B747 first class boundary conditions. The red blocks represent the human bodies and the associated heat sources.

**Table 1.** Boundary conditions

| Boundary | N-S solver | Condition on Energy solver | Species (CO<sub>2</sub>) solver |
| --- | --- | --- | --- |
| **Inlet Jet** | Constant horizontal velocity<br>(0.5 m/s, 10% turbulent intensity) | Constant temperature (18 C) | Zero gradient |
| **Outlet** |  | Zero gradient |  |
| **Human nose** | Constant velocity<br>(0.005 L/s) | Constant temperature (30 C) | Constant concentration<br>(0.00085 mass fraction CO<sub>2</sub>) |
| **Human body** | No slip | Constant flux (55 W) | Zero gradient |
| **Cabin top-bottom walls** | No slip | Insulated | Zero gradient |
| **Cabin side walls** | No slip | Constant temperature (22.8 C) | Zero gradient |
| **Cabin windows** | No slip | Constant temperature (16 C) | Zero gradient |

Human body can be modeled either as a constant temperature boundary or a constant heat source. It was earlier modeled as a constant temperature boundary when the section across human head and torso was considered. But as a result of the absence of human torso from the new section, it was changed to a constant hear source, in the area corresponding to the human torso. Heat source was provided only for the torso region and not for the legs, which was supported by the literature. Human noses were modeled as CO2 sources in the problem. Circles of radius 2 cm were created in the cell zone, with their centers lying at the nose locations, and these were treated as noses.

Four different meshes were created, as shown in figure 4. After the grid convergence study, the finest mesh of size 0.005 m was selected. All subsequent simulations were carried out using this mesh size.

**Figure 4-a.**
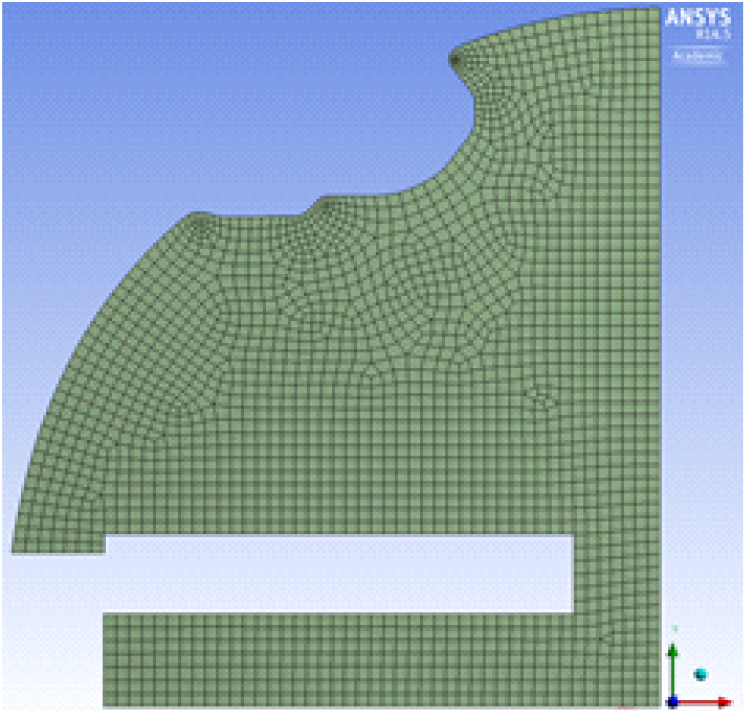
Global size 0.04 m

**Figure 4-b.**
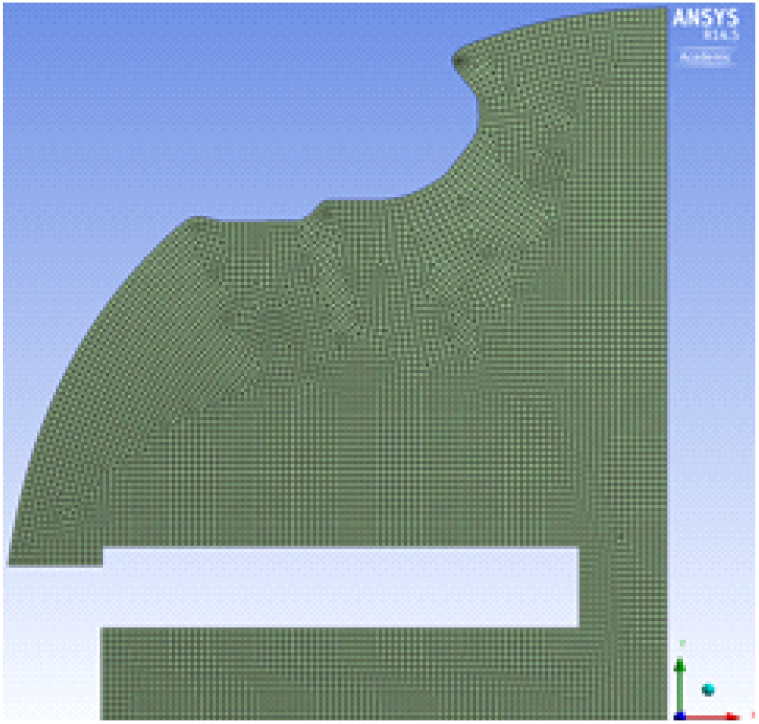
Global size 0.02 m

**Figure 4-c.**
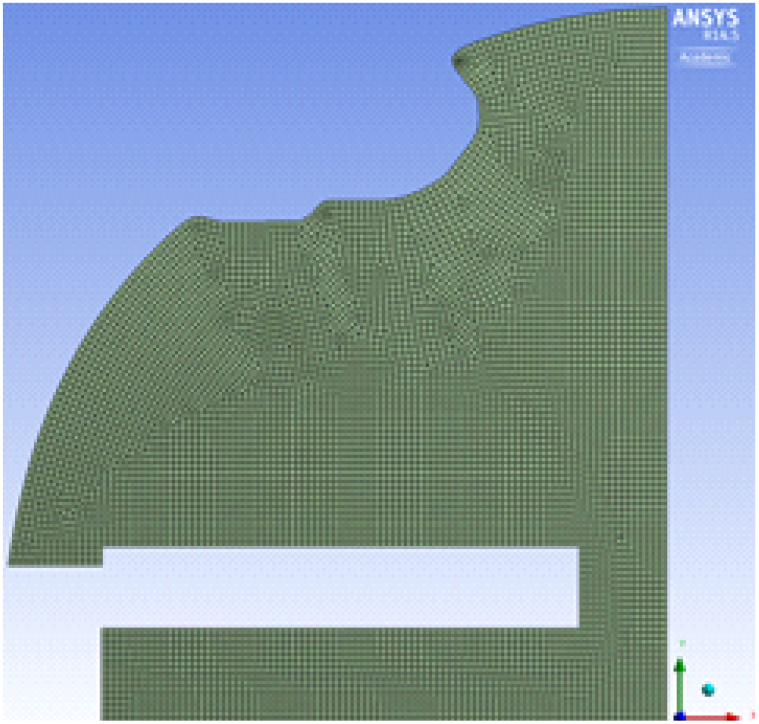
Global size 0.01 m

**Figure 4-d.**
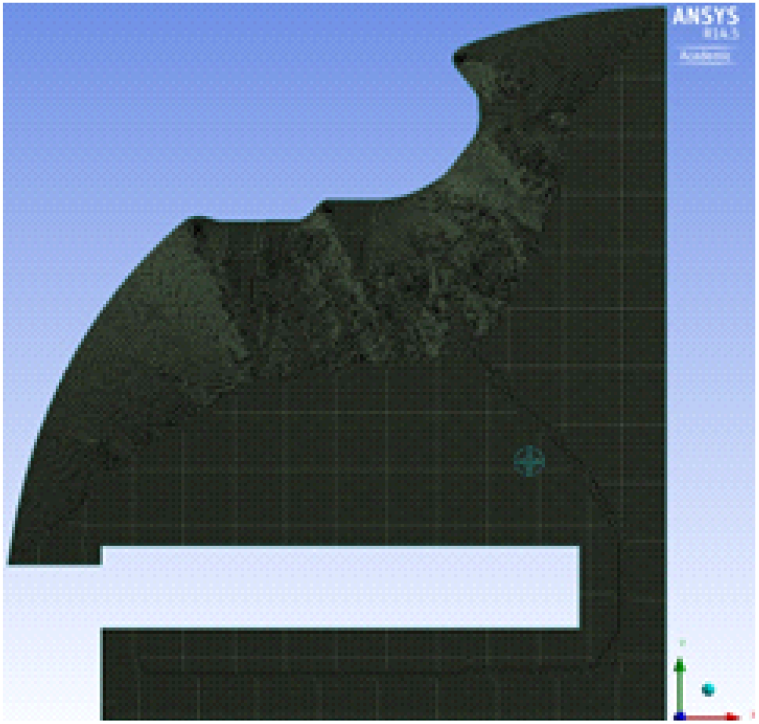
Global size 0.005 m

## NUMERICAL FORMULATION

As a result of the absence of any transient elements from the problem, a 2D steady state simulation was performed using ANSYS Fluent. SIMPLE scheme was used for pressure-velocity coupling. SIMPLE is useful for the problem at hand, as when a steady-state problem is solved iteratively, it is not necessary to fully resolve the linear pressure-velocity coupling, as the changes between consecutive solution are no longer small.

For the SIMPLE algorithm, if the momentum equation is solved with a guessed pressure field *p*\*, the resulting face flux 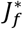 computed from the equation

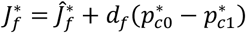

does not satisfy the continuity equation, a correction 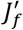 is added to the face flux 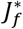 so that corrected flux *J_f_*

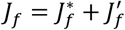

satisfies the continuity equation.

The SIMPLE algorithm postulates that 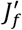 can be written as

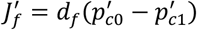

where *p*′ is the pressure correction.

The flux correction equation is then substituted into the discrete continuity equation to obtain the discrete equation for pressure correction

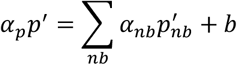

where the source term b is the net flow rate into the cell.

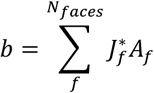

The pressure correction is solved using the algebraic multigrid (AMG) method. Once a solution is obtained, the cell pressure and face flux are corrected using

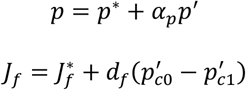

Here *α_p_* is the underrelaxation factor for pressure. The corrected flux, *J_f_* satisfies the discrete continuity equation identically during each iteration.

RANS standard k-*∊* model was used for air flow modeling. The standard k-*∊* model is the most widely used RANS model and was the most preferred model in literature in airplane cabin flow modeling. Hence, we decided to use this model.

Transport equations for the model are as follows:

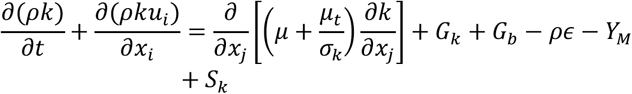

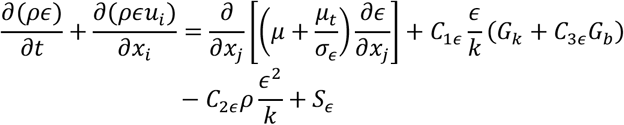

In these equations *G_k_* represents the generation of turbulent kinetic energy due to the mean velocity gradients, *G_b_* is the generation of turbulent kinetic energy due to buoyancy, *Y_M_* represents the contribution of fluctuating dilation in compressible turbulence to overall fluctuation rate, *C*_1*∊*_, *C*_2*∊*_ and *C*_3*∊*_ are constants, *σ_k_* and *σ_∊_* are turbulent Prandtl numbers for k and *∊* respectively, and *S_k_* and *σ*_*∊*_ are user-define source terms.

The turbulent viscosity, *μ_t_*, is computed from k and *∊*, as follows:

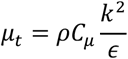

where *C_μ_* is a constant.

The model constants *C*_1*∊*_, *C*_2*∊*_, *C*_3*∊*_, *σ_k_* and *σ*_*∊*_, have the following default values:

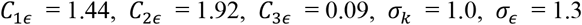

Temperature distribution in the cabin was modeled by the default energy equation in ANSYS Fluent. The species transport module was used to model CO2 transport with CO2 and air being treated as two different species.

Final aim was to rank the seats in Airbus and Boeing sections using the simulation results for velocity, temperature, and CO2 mass concentration. Following ranking scheme was developed:

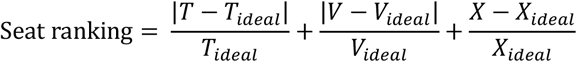

where,

*T_ideal_* – Ideal temperature = 294 K
*V_ideal_* – Ideal velocity = 0.2 m/s
*X_ideal_* – Ideal mass concentration of CO2 = 1.2 × 10^−3^

An absolute value was not used for CO2 concentration, because lower the value of CO2, the better. The ideal temperature and velocity values were obtained from the ASHRAE handbook detailing standards for ambient environmental conditions for the human comfort [22].

As seen from the ranking scheme, closer the values are to the ideal the value, better the seat, and better the rank. Hence, seat with the lowest rank is the best seat.

## RESULTS AND DISCUSSION

A grid-converge study was carried out with mesh details as shown in Table 2. Meshes 1, 2, and 3 are shown in Fig. 4-a to 4-c. Airbus A380 Business Class section was used for this study. This section has two seats and is located on the upper deck of A380 (refer Fig. 14-c). 2-D line plots at the nose-level of passengers for x-Velocity, mass fraction of CO2, and total temperature are shown in Figs. 5 through 7 respectively. The results become invariant as for Mesh 3 and beyond. Mesh 4 is also able to capture the thermal boundary-layer developed at the cold window on the left side of the aforementioned section.

**Table 2.** Mesh-convergence study

| Mesh | Global size (m) | Number of nodes | Number of elements |
| --- | --- | --- | --- |
| 1 | 0.04 | 2130 | 1981 |
| 2 | 0.02 | 7832 | 7556 |
| 3 | 0.01 | 30435 | 29900 |
| 4 | 0.075 | 53570 | 52856 |

**Figure 5.**
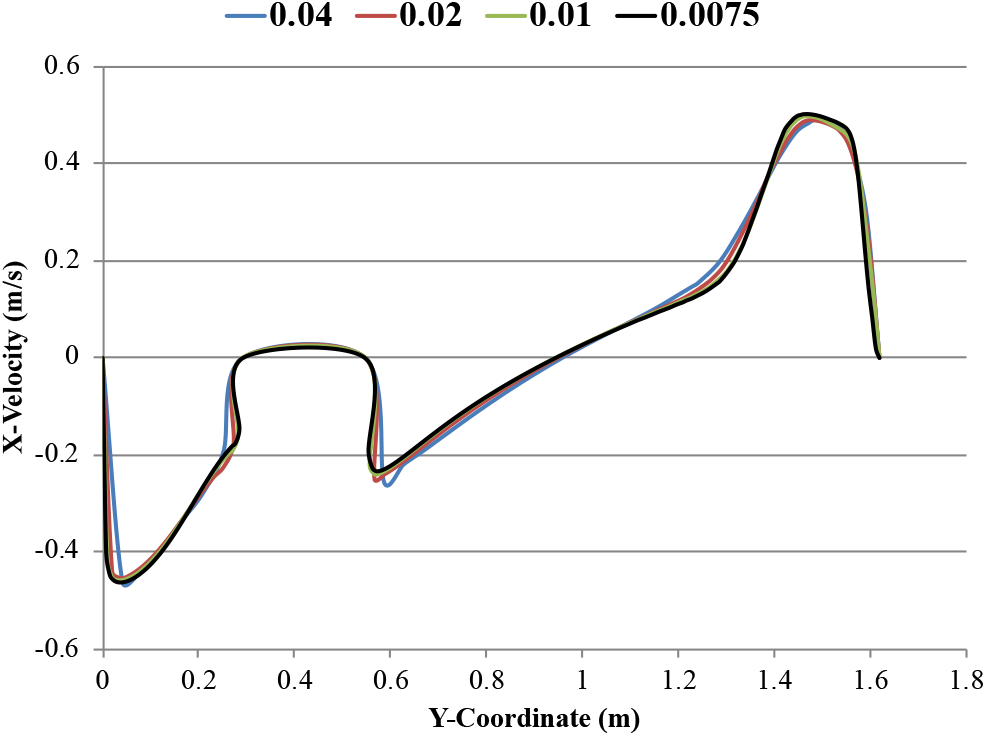
Mesh-convergence: x-velocity

Authors of this paper felt the need to refine the mesh further to a global size of 0.005 m to account for any changes which the other sections, first and economy, might bring in due to a larger size and/or more passengers. All the remaining results have been obtained using a global element size of 0.005 m. Simulations were carried out for the following six sections:

1. Airbus A380 First, Business and Economy class (3 sections)
2. Boeing B747 First, Business and Economy class (3 sections)

Contour and line plots shown in Figs. 8 through 13 are for the first class section of A380 and B747. The Airbus section has two seats and the Boeing one has three seats. Figure 8 shows the steady state contours for the velocity magnitude for the first class section of A380 (Fig. 8-a) and B747 (Fig. 8-b). The corresponding line plots for the velocity magnitude at the nose-level are shown in Fig. 9. Two big eddies are seen in A380 while a single large eddy is seen in B747. Velocity magnitude plots at the nose-level for first class sections of A380 and B747 reveal that the velocity magnitude in B747 follows a monotonic trend while moving from the center of the cabin (aisle) to the window. This is due to the presence of a single large eddy seen in B747 as opposed to A380, that has an unpredictable trend in velocity magnitude at the nose-level.

**Figure 6.**
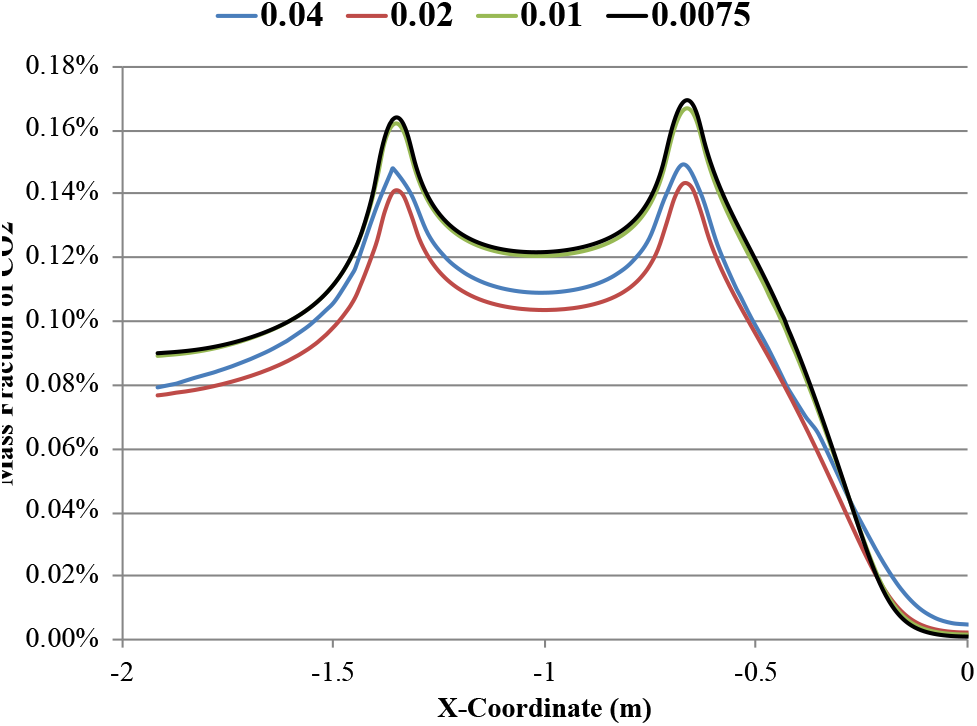
Mesh-convergence: mass fraction of CO_2_

**Figure 7.**
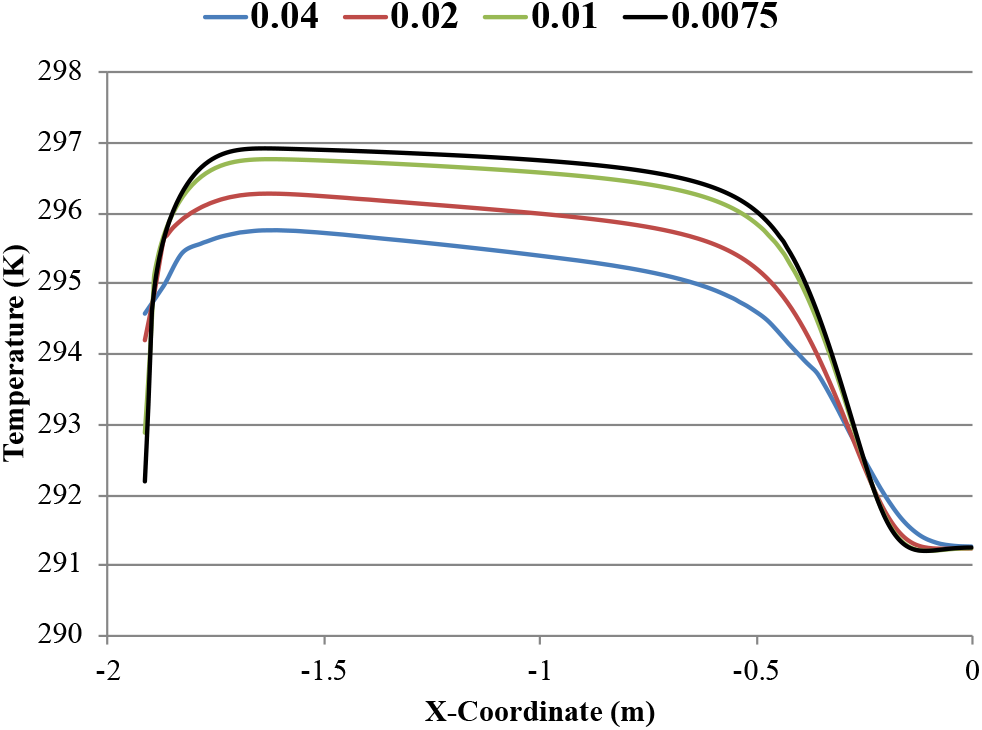
Mesh-convergence: Total temperature

Figure 8.

Steady state contours of velocity magnitude

**Figure 8-a.**
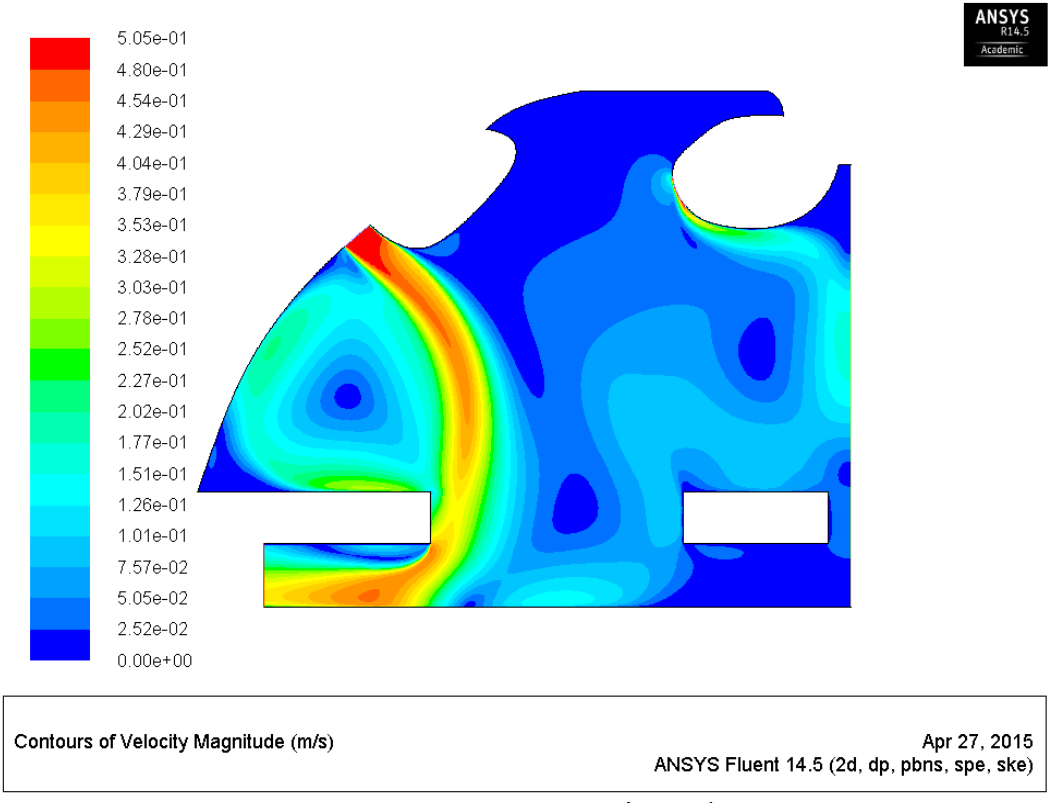
A380 First Class.

**Figure 8-b.**
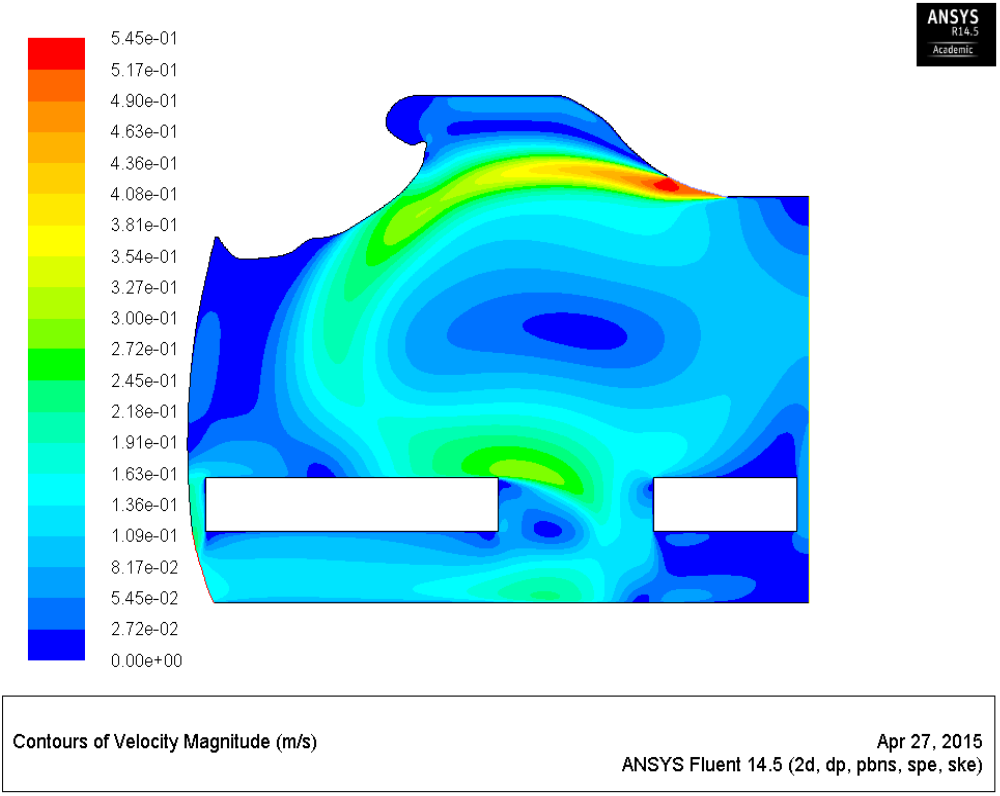
B747 First Class.

Figure 10 shows the steady state contours for the mass concentration of CO2 for the first class section of A380 (Fig. 10-a) and B747 (Fig. 10-b). The corresponding line plots for the mass concentration of CO2 at the nose-level are shown in Fig. 11. Two big eddies are seen in A380 while a single large eddy is seen in B747. The CO2 released by the two passengers in A380 seems to circulate around them as opposed to that in B747 where the CO2 released by three passengers seems to be swept away more efficiently towards the outlet. The resulting higher concentration of CO2 at the nose-level of passengers in A380 can be seen in Fig. 11-a while that in B747 is shown in Fig. 11-b.

Figure 9.

Nose-level line plot of steady state velocity magnitude

**Figure 9-a.**
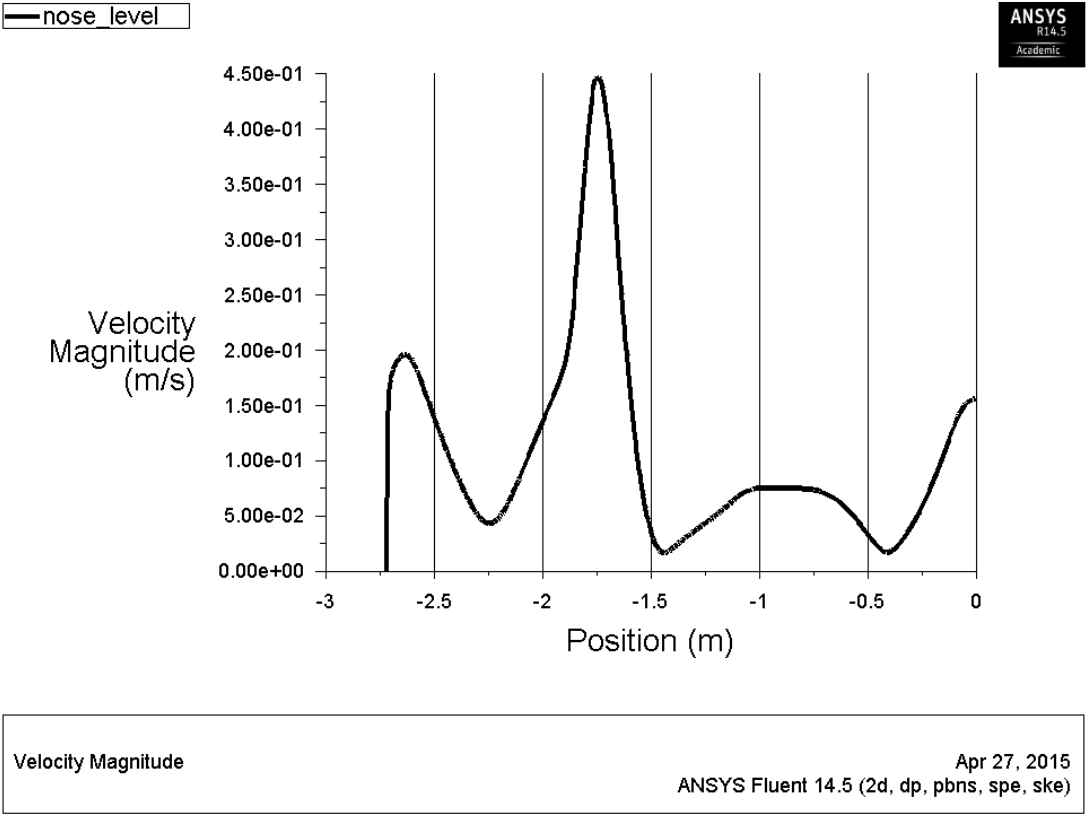
A380 First Class.

**Figure 9-b.**
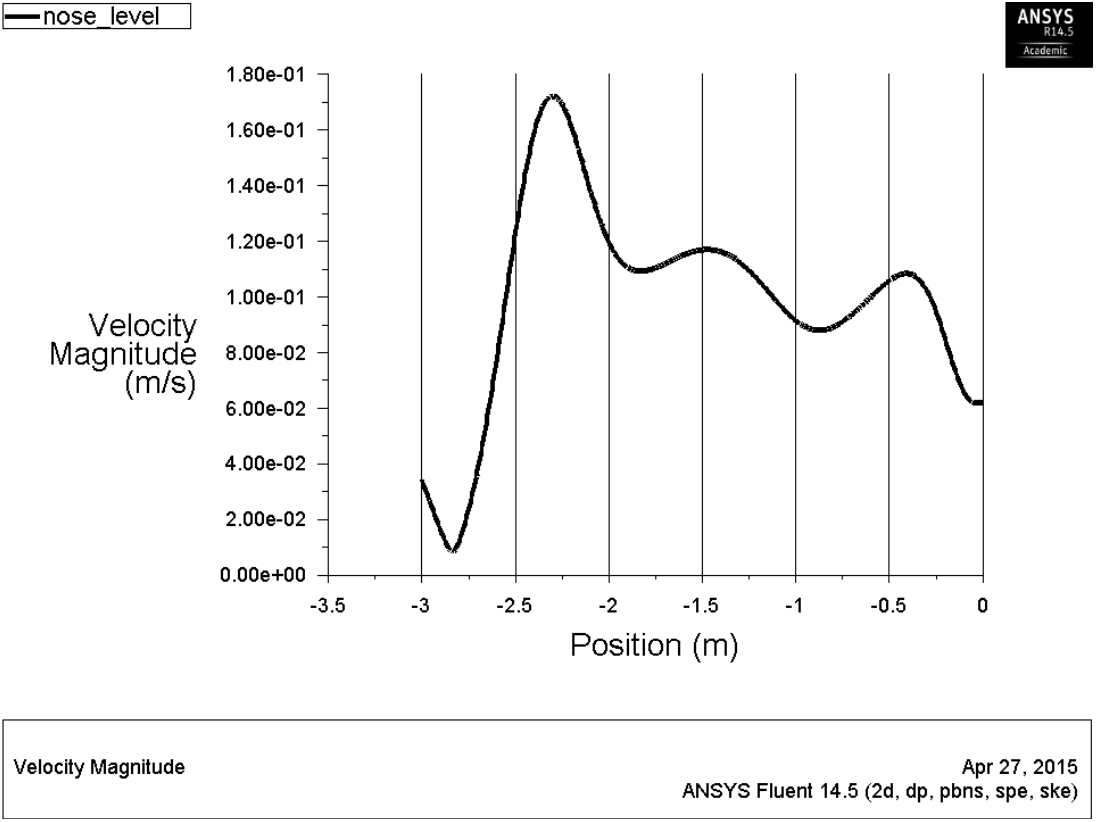
B747 First Class.

Figure 10.

Steady state contours of mass fraction of CO_2_

**Figure 10-a.**
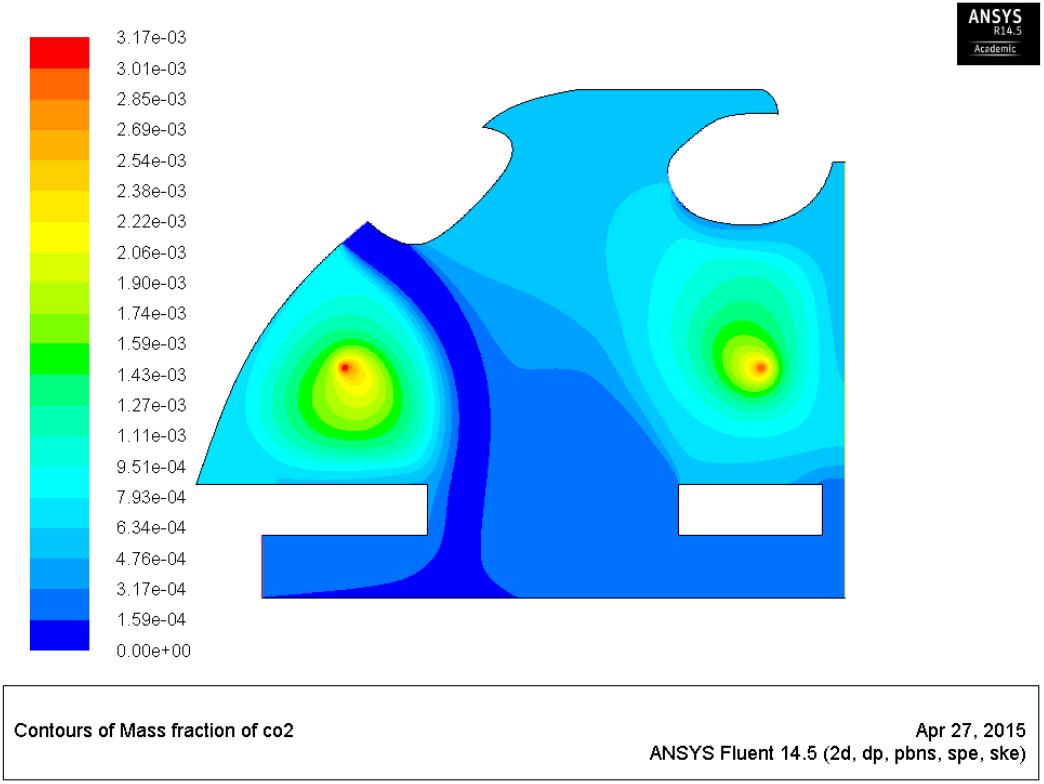
A380 First Class.

**Figure 10-b.**
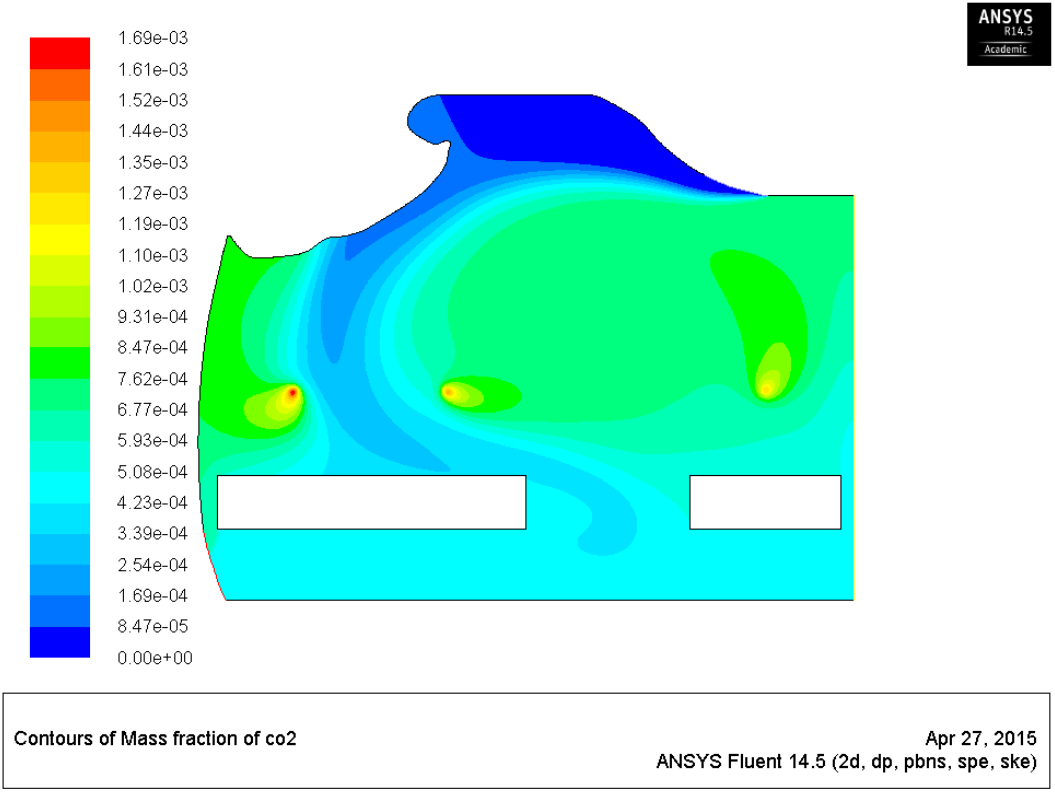
B747 First Class.

Figure 11.

Nose-level line plot of steady state mass fraction of CO_2_

**Figure 11-a.**
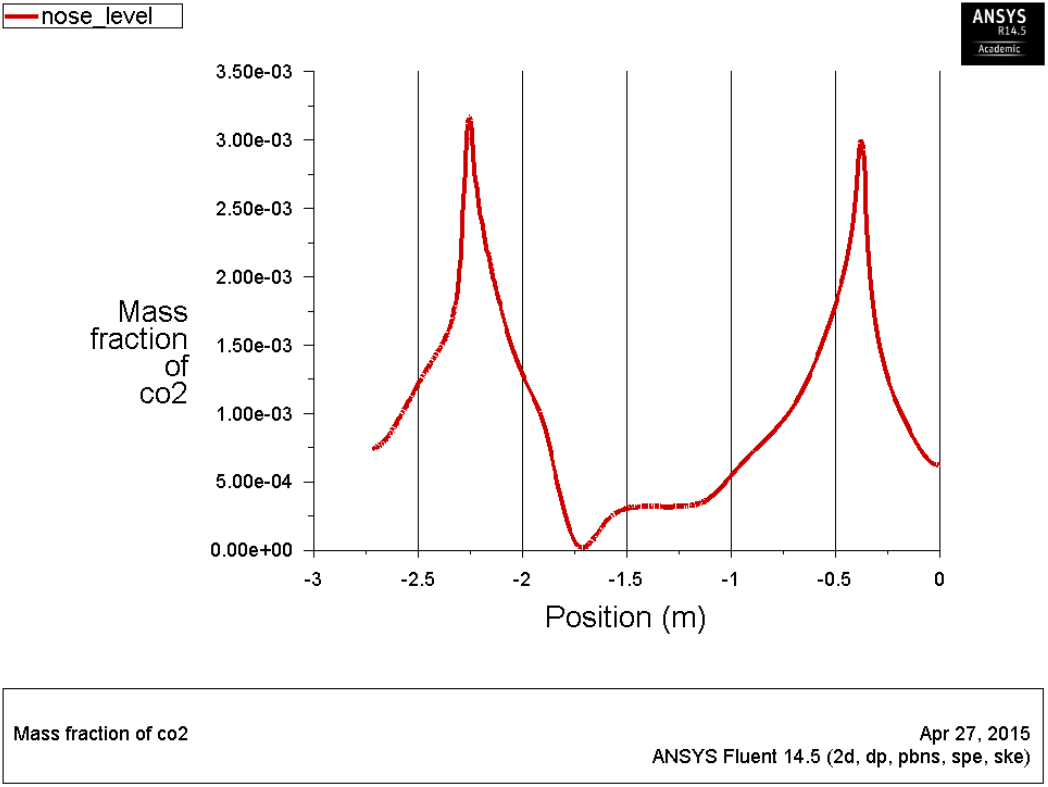
A380 First Class.

**Figure 11-b.**
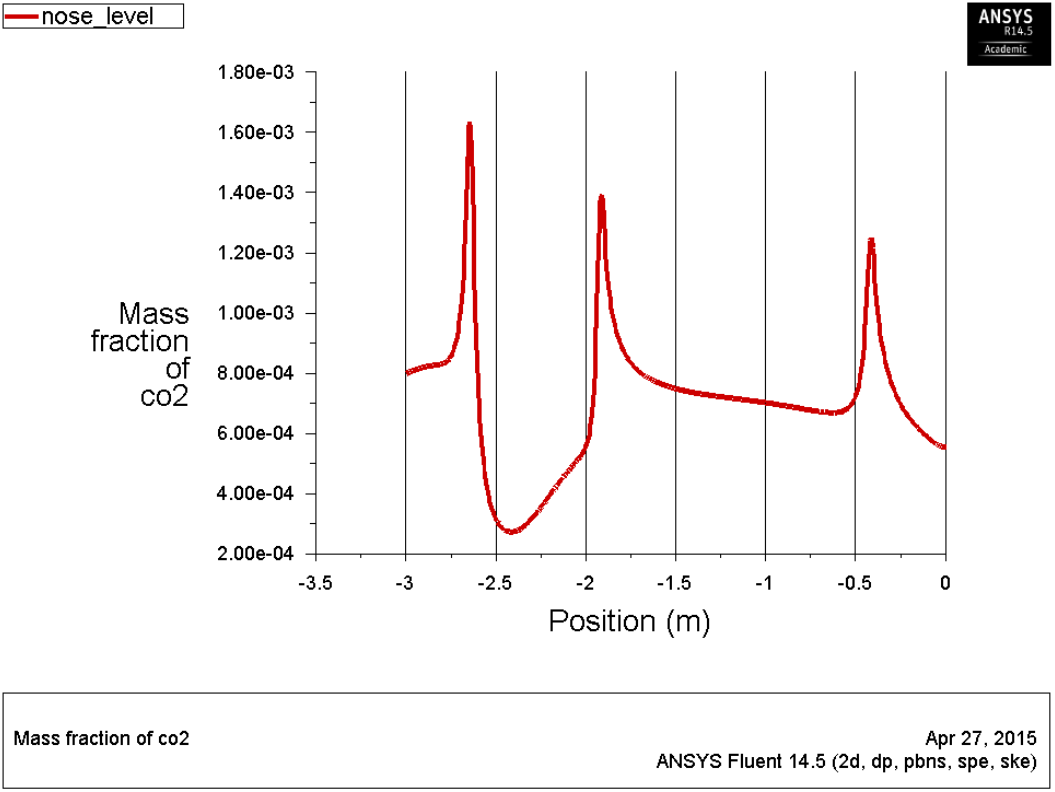
B747 First Class.

Figure 12 shows the steady state contours for the total temperature for the first class section of A380 (Fig. 12-a) and B747 (Fig. 12-b). The corresponding line plots for the total temperature at the nose-level are shown in Fig. 13. A large warmer zone can be seen near the window of A380 while the temperature in A747 seems to be pretty uniform for the major part of the cabin. This can be further illustrated by observing the line plots of Fig. 13.

Figure 12.

Steady state contours of total temperature

**Figure 12-a.**
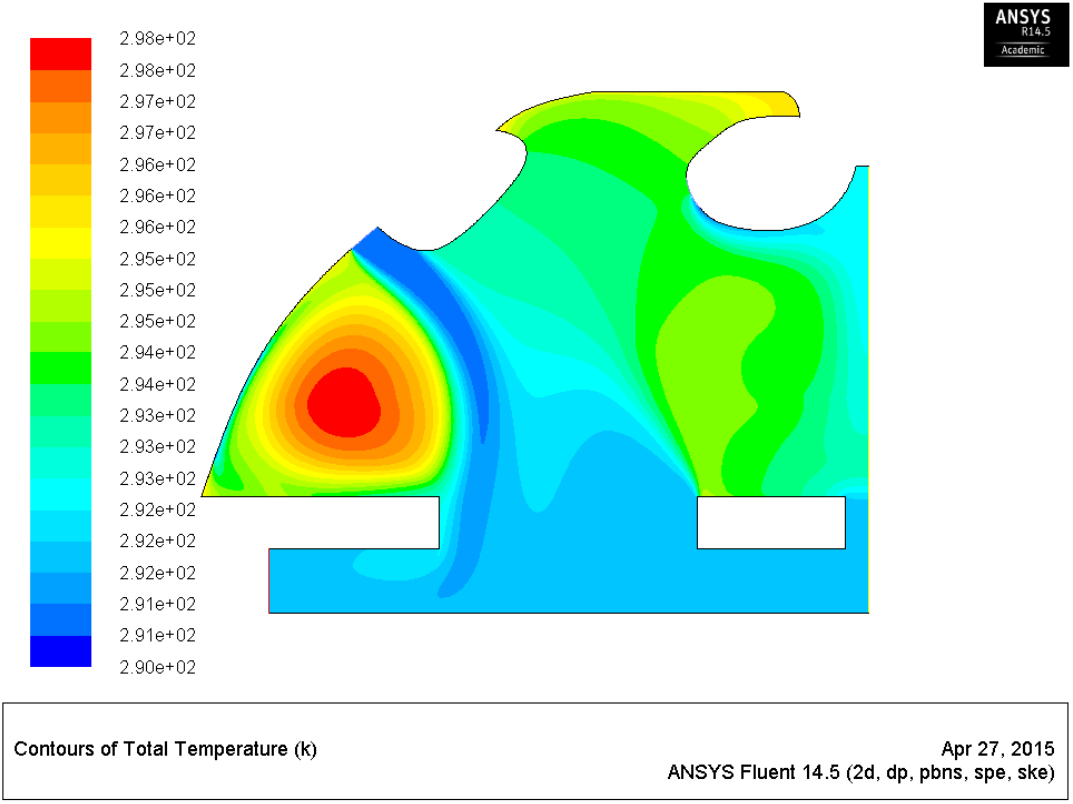
A380 First Class.

**Figure 12-b.**
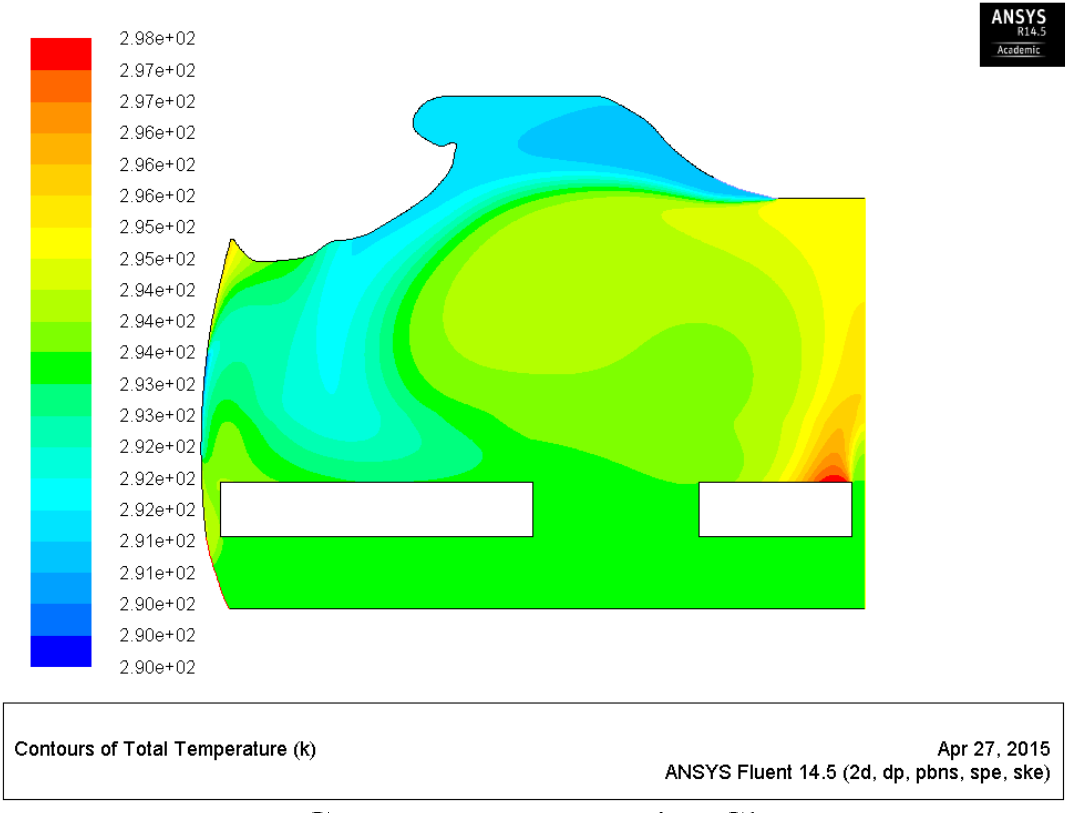
B747 First Class.

Figure 13.

Nose-level line plot of steady state total temperature

**Figure 13-a.**
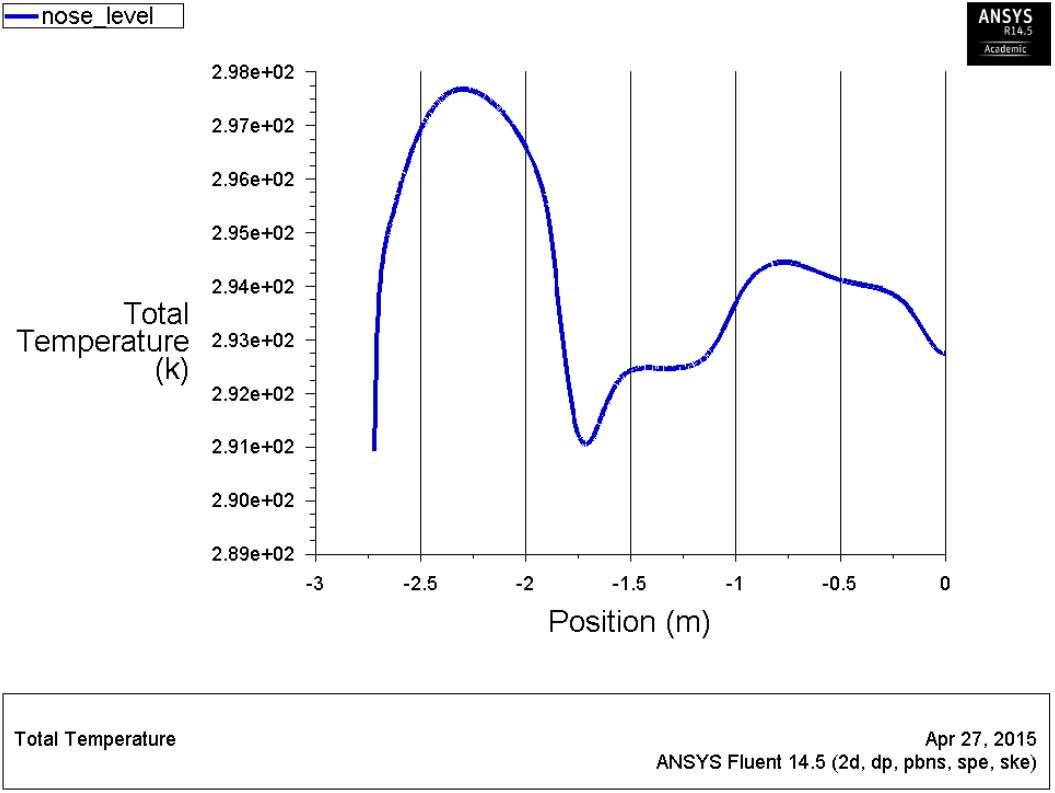
A380 First Class.

**Figure 13-b.**
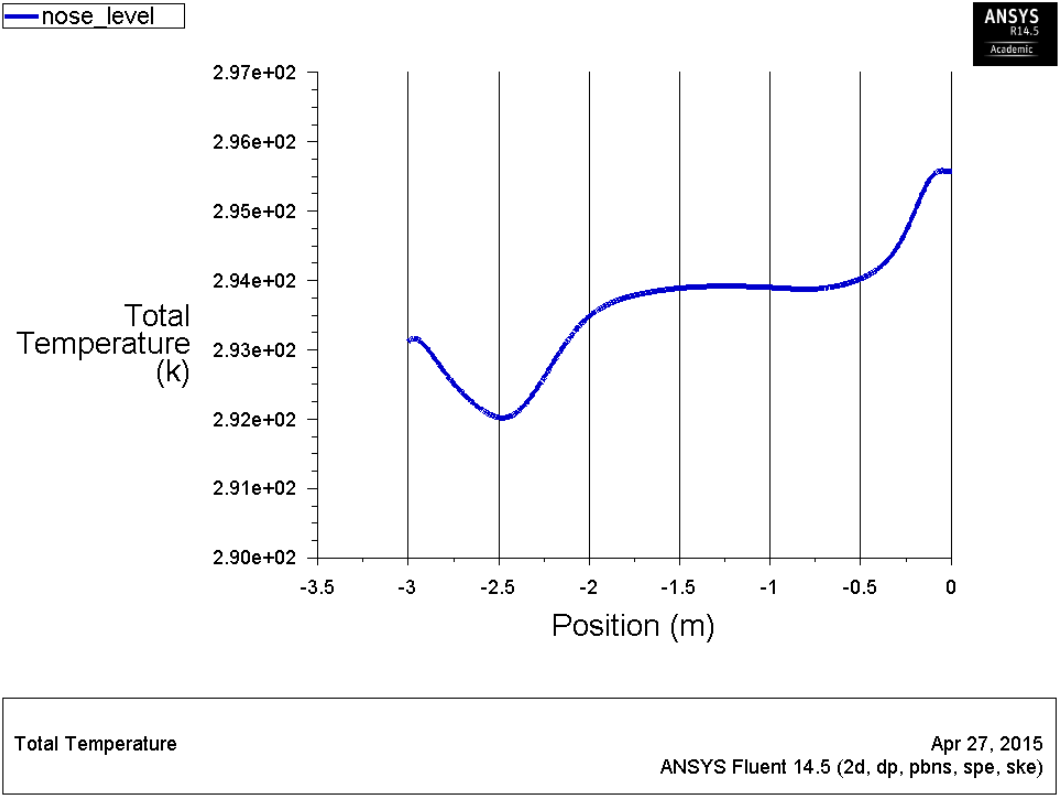
B747 First Class.

Ranking scheme described in the previous section helps to better combine this data from Figs. 8 through 13 for the first class sections of A380 and B747 in the form of a figure of merit like quantity based on the equation describing the seat ranking. Lower the value of the ranking quantity, better the seat, and in turn higher the rank. The ranking obtained using this approach is schematically shown in Fig. 14-a and Fig. 14-b. As can be seen in these two figures, A380’s first class seats tie as per the ranking scheme while B747’s center seat is the most superior when compared to the window and second to window seats. It should be noted that mirror image of the shown ranking exists on the right half of the cabin sections.

Figure 14.

Airbus vs. Boeing: Seat rankings for different classes [23]

**FIGURE 14-a.**
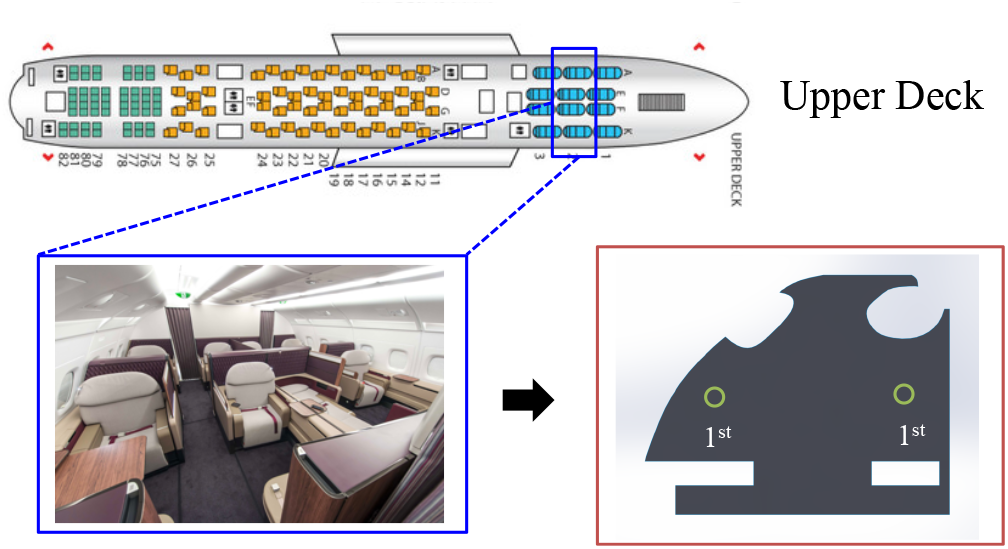
Airbus A380 First

**FIGURE 14-b.**
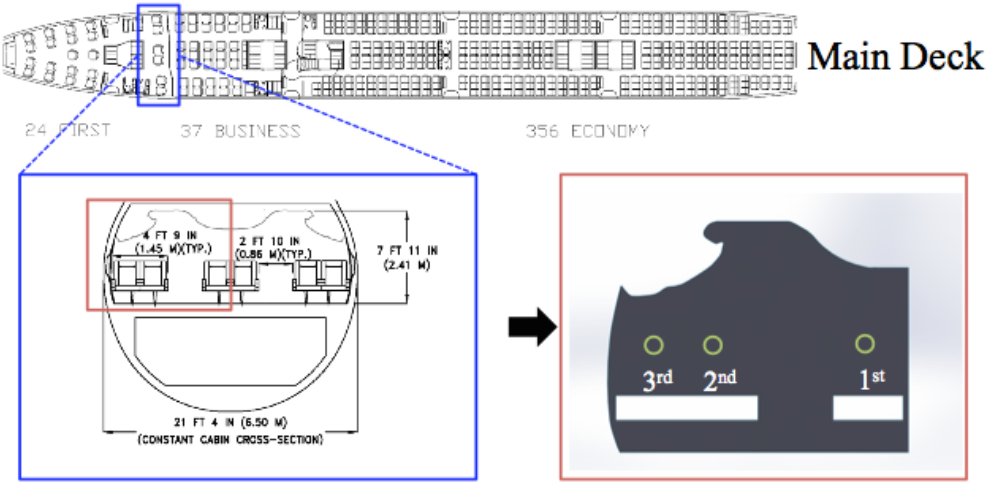
Boeing B747 First

**FIGURE 14-c.**
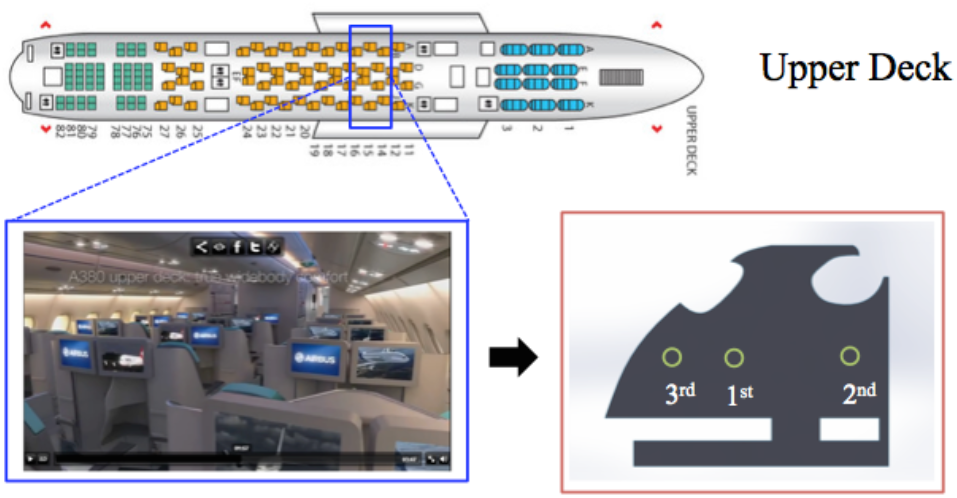
Airbus A380 Business

**FIGURE 14-d.**
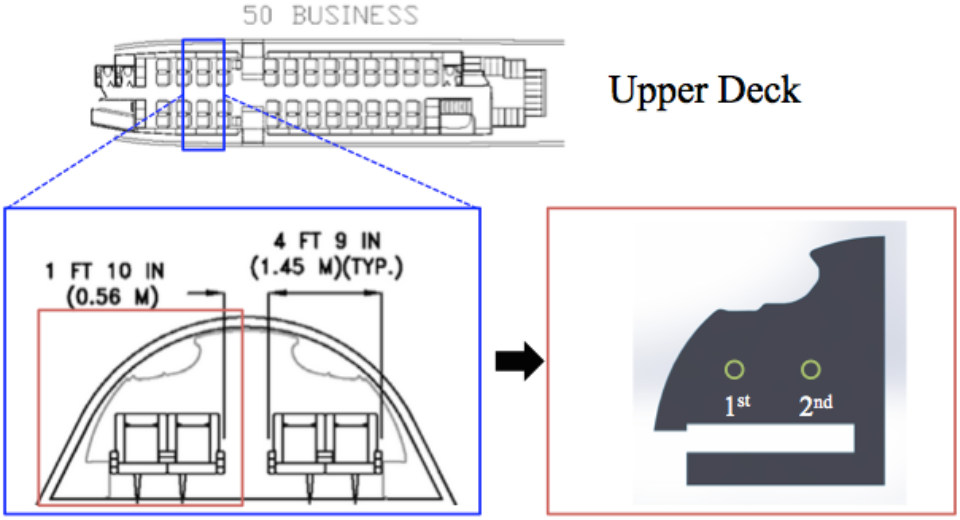
Boeing B747 Business

**FIGURE 14-e.**
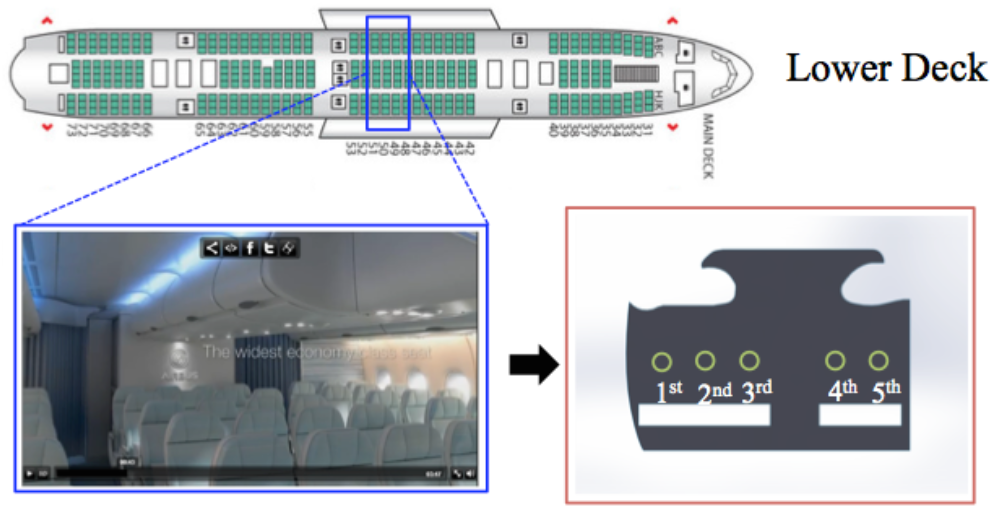
Airbus A380 Economy

**FIGURE 14-f.**
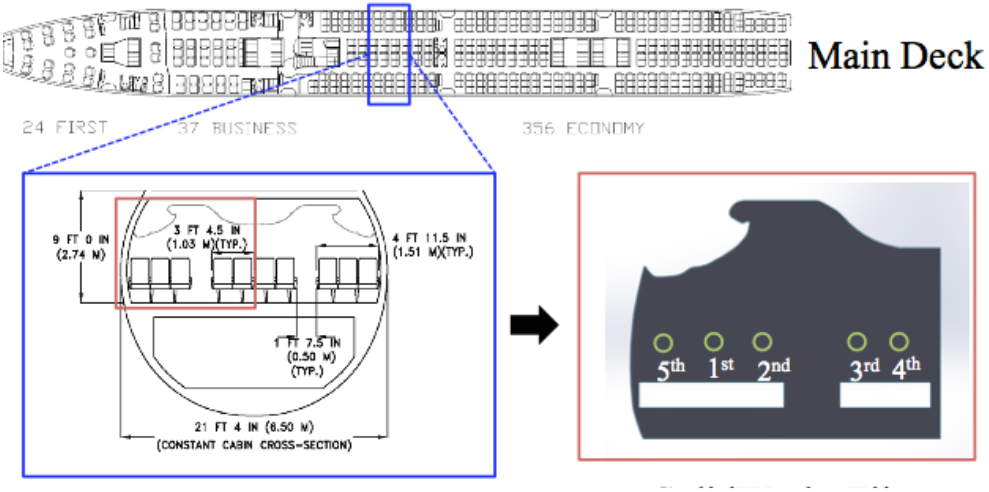
Boeing B747 Economy

Rankings obtained using a similar procedure for the remaining four sections of interest are shown in Figures 14-c through 14-f. As with the first class sections, even the business and economy sections of A380 and B747 have dis-similar rankings for seats occupying same relative positions. In other words, a window seat in the economy class of A380 would be most preferred but its counterpart, an economy window for B747, would be least preferred. Other results have been presented in the Appendix.

### Limitations

The study assumes 2D flow pattern for the ease of caluculations. This is backed by previous studies and is quite correct for the central portion of the aircraft. However, authors do recognize that there can 3D motion of air and thereby the pollutants released orally or nasally into the air (viz., CO2 and viruses such as coronavirus) can travel in the longitudinal directions (i.e., along the length of the aircraft). This is partiularly important to consider at the ends of the aircraft as the fluid flow at the ends will certainly not be 2D. The BCs were based on the data available in the public doamin or by making conservative assumptions. These may not accuratley describe the air duct positions in the studied aircrafts. As a simplifying assumption that also adds to the factor of safety of the model predictions, coronavirus was assumed to be present at places where released CO_2_ can be found. The released CO_2_ and the associated coronavirus concentration has been modeled as a continnum where as the actual coronavirus particles would be in a discrete phase. Modeling coronavirus as a discrete phase suspended in air will be prohibitively expensive with regards to the computational resources.

These rankings are based on the scheme presented by the authors and the section views and BCs as available in the public domain. Many simplifying assumptions are associated with this study and the actual ranking of the seats can be different based on how many of these assumptions fail in real life and personal preference of the traveler.

## CONCLUSIONS

Many factors are considered during the decision making process of purchasing a seat on a plane. Features such as costumer service, financial means, and inherent bias often drive the decision making process. However, these factors are not considered in our conclusions about the ideal airplane seat. Furthermore, personal preference plays a significant role when choosing a seat. For example, a particular individual may prefer to sit by the window so he/she can see the view, or sit by the aisle so he/she can stretch his/her legs. Also the ideal enviornmental conditions used in the ranking scheme described previously are an average and not the same for each individual. Some passengers may prefer a hotter ambient temperature and other slightly cooler. For all these reasons we will present our findings in a way that will no make any assumptions about the passenger, but instead provide the reader with all the information necessary to make a more educated purchase with regard to their own personal prefernces. This information is as shown in Tables 3 through 5.

**Table 3.** First Class

|  | Airbus | Boeing |
| --- | --- | --- |
| **CO<sub>2</sub> (mass frac)** | 3e-03 | 1e-03 |
| **Temperature** | 25 C | 21 C |
| **Velocity (m/s)** | 0.0437 | 0.108 |

Each of the abovementioed tables contains data for the three sections of the airplane: first class, business class, and economy class. The section is generally chosen based on the financial means of the traveller so it is unnessasary to compare data between classes. Each row of the table contains data for each of the enviornemntal conditions studied in this report. The mass fraction of CO2 that can be qualitatively thought of as the mass fraction of orally or nasally released pollutants, is reported as well as the temperature and air velocity at the seat. The data is reported for the best seat in every section for the Boeing cabin and the Airbus cabin. This enables a comparison across classes between the two planes.

In Table 3 the data for the first class cabins for Airbus and Boeing are shown. We see that the Airbus seat is warmer than the Boeing seat, but has worse circulation. This decreased circulation is confirmed by the lower air velocity and increased mass fraction of CO2. The Airbus seat will be warm, but with the potential to be “stuffy”. Conversley the Boeing seat, located in the middle on the aisle side, will be cooler and breezy.

Table 4 shows the data for the business class sections of both the planes. Here the effects are reversed with respect to the first class cabin. We see that the Airbus seat is colder, but offers better circulation than the Boeing seat. The Airbus seat is located in the side bank of the seats on the aisle side and the Boeing seat, that is warmer compared to the Airbus seat and with worse circulation, is located next to the window.

**Table 4.** Business class

|  | Airbus | Boeing |
| --- | --- | --- |
| **CO<sub>2</sub> (mass frac)** | 8e-04 | 1e-03 |
| **Temperature** | 21 C | 24 C |
| **Velocity (m/s)** | 0.33 | 0.048 |

The results for the economy class of both planes are shown in Table 5. We see that the best seat for the Airbus cabin is located next to the window while the best seat for the Boeing cabin is the middle seat in the side bank of the seats. The Airbus seat has a higher temperature, lower CO2 concentration, and lower air velocity. Unlike the other sections were the tradeof for a warmer seat was worse circulation, we conclude that the Airbus economy best seat is both warm and with good circulation. The Boeing seat performs less well in all these areas.

**Table 5.** Economy class

|  | Airbus | Boeing |
| --- | --- | --- |
| **CO<sub>2</sub> (mass frac)** | 1e-03 | 1e-03 |
| **Temperature** | 22 C | 20 C |
| **Velocity (m/s)** | 0.086 | 0.15 |

The findings from this preliminary multiphysics computational model may be used by the general public to decide which seat to occupy for their next intercontinental flight. Alternatively, the commercial airliners can use such a model to plan the occupancy of the aircraft on long-duration intercontinental flights (viz., Airbus A380 and Boeing B747).

## Data Availability

All relevant data can be reproduced by running the multiphysics simulations, the details for which are mentioned in the manuscript.

## ACKNOWLEDGMENTS

1. Dr. S. Singh, Associate Teaching Professor, Carnegie Mellon University for his guidance and brainstorming activities with the authors.
2. Pittsburgh Supercomputing Center (PSC) for computational resources.

## AUTHOR CONTRIBUTIONS

All authors contributed equally to the design and performance of the research, data analysis, writing, and editing the manuscript.

## SUPPLEMENTARY MATERIAL

## Appendix A

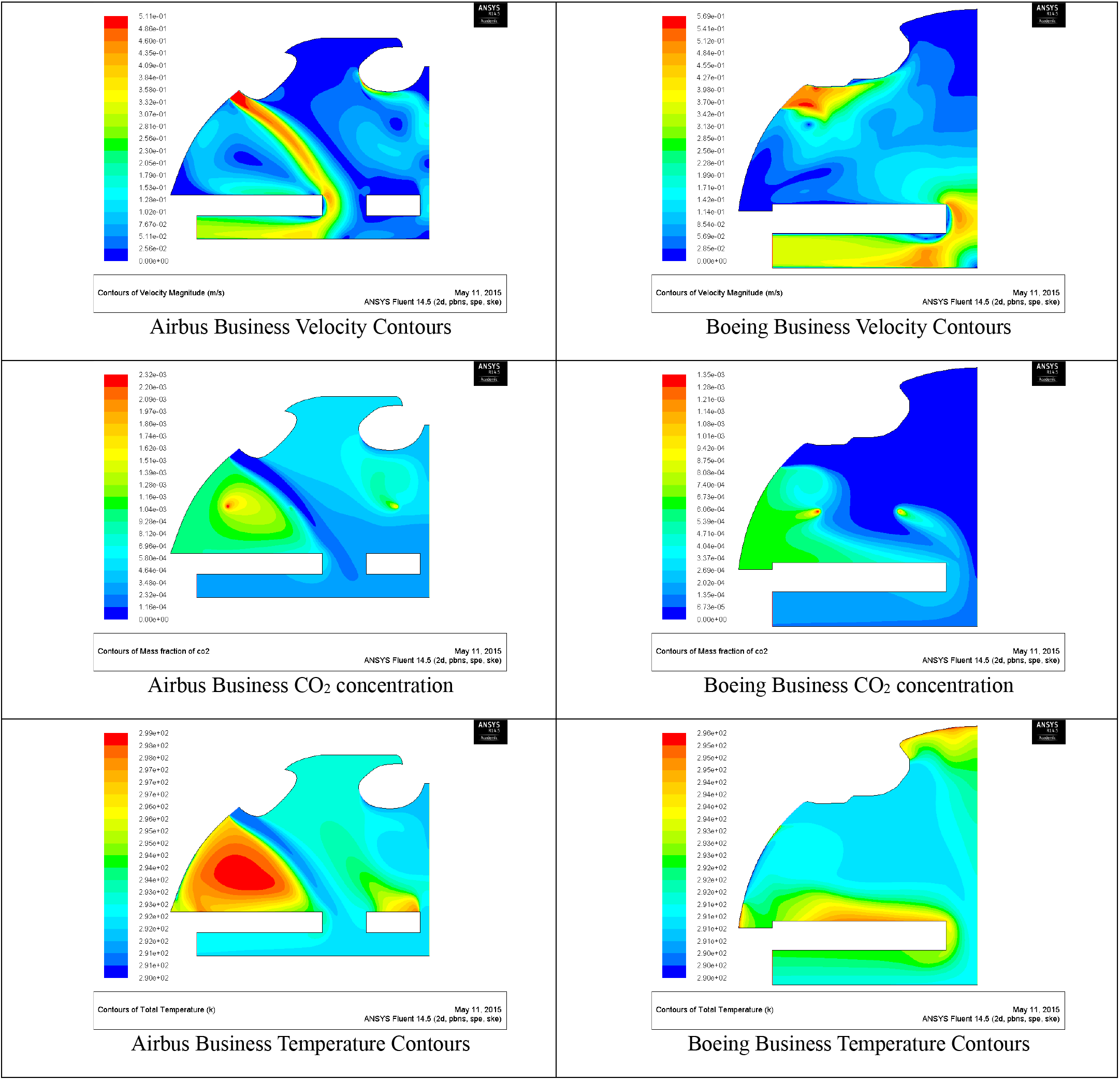
Graphical Results for Business Class

## Appendix B

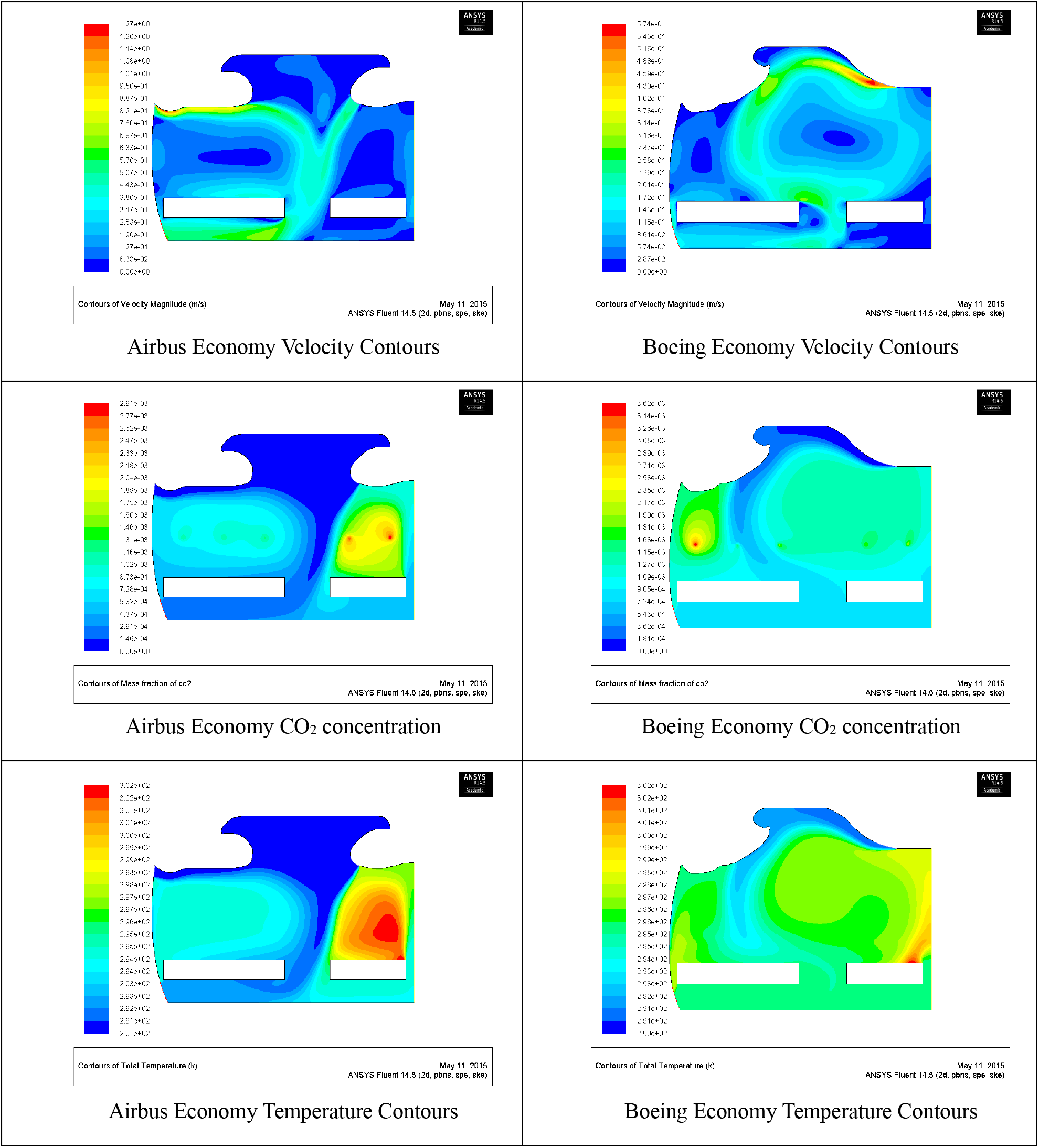
Graphical Results for Economy Class

## REFERENCES

1. Anderson, M., Mckee, M. and Mossialos, E., 2020. Developing a sustainable exit strategy for COVID-19: health, economic and public policy implications. Journal of the Royal Society of Medicine, 113(5), pp.176–178. p.248–253

2. Wilbur, M., Ayman, A., Ouyang, A., Poon, V., Kabir, R., Vadali, A., Pugliese, P., Freudberg, D., Laszka, A. and Dubey, A., 2020. Impact of COVID-19 on Public Transit Accessibility and Ridership. arXiv preprint arXiv:2008.02413.

3. Ferris, R. 2020. Why Hertz landed in bankruptcy court when its rivals didn’t (Available online: https://www.cnbc.com/2020/08/17/why-hertz-landed-inbankruptcy-court-when-its-rivals-didnt.html) [the easiest access to this source is via the URL]

4. Singh, A., Hosni, M.H. and Horstman, R.H., 2002. Numerical simulation of airflow in an aircraft cabin section/Discussion. ASHRAE Transactions, 108, p.1005.

5. Zhang T, Yin S, Wang S. “An under-aisle air distribution system facilitating humidification of commercial aircraft cabins”. Building and Environment 2010; 45(4):907–15.

6. Zhang, T., P. Li, and S. Wang. “A personal air distribution system with air terminals embedded in chair armrests on commercial airplanes”. Building and Environment 47(1):89–99 2012a.

7. Yan W, Zhang Y, Sun Y, Li D. “Experimental and CFD study of unsteady airborne pollutant transport within an aircraft cabin mock-up”. Build Environ 2009; 44: 34–43.

8. Wu, Chaofan and Ahmed, Noor A. “Numerical Study of Transient Aircraft Cabin Flow field with Unsteady Air Supply” Journal of Aircraft, Vol. 48, No. 6 (2011), pp. 1994–2001.

9. Liu, W., Mazumdar, S., Zhang, Z., Poussou, S.B., Liu, J., Lin, C.-H., Chen, Q. “State-of-the-art methods for studying air distributions in commercial airliner cabins”. Building and Environment 47, 5–12. 2012

10. Liu W, Wen J, Chao J, Yin W, Shen C, Lai D,. “Accurate and high-resolution boundary conditions and flow fields in the first-class cabin of an MD-82 commercial airliner”. Atmospheric Environment. 2012; 56:33–44.

11. Wang A, Zhang Y, Sun Y, Wang X. “Experimental study of ventilation effectiveness and air velocity distribution in an aircraft cabin mockup”. Build Environ 2008;43(3):337–43.

12. Garner R, Wong K, Ericson S. “CFD validation for contaminant transport in aircraft cabin ventilation flow fields”. Proceedings of Annual SAFE Symposium 2003.

13. Bosbach, Johannes and Kühn, Matthias and Rütten, Markus and Wagner, Claus (2006) “Mixed Convection in a Full Scale Aircraft Cabin Mock-Up”. ICAS 2006 – 25th International Congress of the Aeronautical Sciences (Paper No. 715), pp. 1–9. ISBN 0-9533991-7-6.

14. Perella, P., Tabarra, M., Hataysal, E., Pournasr, A. and Renfrew, I., 2020. Minimising exposure to droplet and aerosolised pathogens: A computational fluid dynamics study. medRxiv.

15. Adwibowo, A., 2020. Computational fluid dynamic (CFD), air flow-droplet dispersion, and indoor CO2 analysis for healthy public space configuration to comply with COVID 19 protocol. medRxiv.

16. Vuorinen, V., Aarnio, M., Alava, M., Alopaeus, V., Atanasova, N., Auvinen, M., Balasubramanian, N., Bordbar, H., Erästö, P., Grande, R. and Hayward, N., 2020. Modeling aerosol transport and virus exposure with numerical simulations in relation to SARS-CoV-2 transmission by inhalation indoors. Safety Science, p.104866.

17. Li, Y., Qian, H., Hang, J., Chen, X., Hong, L., Liang, P., Li, J., Xiao, S., Wei, J., Liu, L. and Kang, M., 2020. Evidence for probable aerosol transmission of SARS-CoV-2 in a poorly ventilated restaurant. medRxiv.

18. Garbey, M., Joerger, G. and Furr, S., 2020. A Systems Approach to Assess Transport and Diffusion of Hazardous Airborne Particles in a Large Surgical Suite: Potential Impacts on Viral Airborne Transmission. medRxiv.

19. Li, F., Liu, J., Ren, J., Cao, X. and Zhu, Y., 2016. Numerical investigation of airborne contaminant transport under different vortex structures in the aircraft cabin. International Journal of Heat and Mass Transfer, 96, pp.287–295.

20. Bhatia, D. and De Santis, A., 2020. A preliminary numerical investigation of airborne droplet dispersion in aircraft cabins. Open Journal of Fluid Dynamics, 10(3), pp.198–207.

21. Mittal, R., Ni, R. and Seo, J.H., 2020. The flow physics of COVID-19. Journal of fluid Mechanics, 894.

22. ASHRAE. “ASHRAE handbook fundamentals”. Atlanta: American Society of Heating, Refrigerating and Air-Conditioning Engineers; 2009.

23. Official websites of Boeing and Airbus: www.boeing.com and www.airbus.com

